# New universal approach for microplastics detection in tissues retains histology and reveals unprecedented quantities in placental samples

**DOI:** 10.1101/2025.08.25.25334346

**Authors:** Quinten Wouters, Sergey Abakumov, Charlotte Van Der Stukken, Imran Aslam, Iris Van Den Eede, Peter Dedecker, Tim S. Nawrot, Maarten B.J. Roeffaers

## Abstract

**Background:** Micro- and nanoplastics (MNPs) contamination may pose a significant risk to human health. However, their true impact remains underexplored due to substantial limitations of current analytical methods. Traditional techniques like Raman and FTIR microscopy, coupled with filtration, fail to detect smaller MNPs and are prone to external contamination. Likewise, pyrolysis-GC/MS lacks the ability to pinpoint MNP size or location.

**Methods:** This study presents a universal approach for MNPs detection in tissues, validated to mitigate external contamination risks and enable the identification of significantly smaller MNPs. The method preserves histological information, allowing for comprehensive spatial analysis, including assesment of local DNA damage using the γ-H2AX histone.

**Findings:** Applying this method to human placenta samples revealed orders of magnitude higher MNP loads than previously reported, with quantities ranging from thousands to millions per cm³, far exceeding current reports of fewer than 1 MNP per gram or cm^3^ of tissue. Importantly, within the observed concentration range, we found a positive association between MNP load and placental DNA damage.

**Interpretation:** Our findings show that that the prevalence of MNPs in biological tissues has been substantially underestimated, as the smallest and potentially most harmful MNPs go undetected with traditional methods. Furthermore, we found that the concentations were linked with genotoxic effects in the placenta. This novel analytical workflow represents a significant advancement in MNPs research and provides crucial insights into their impact on human health.

**Research in context:** *Evidence before this study:* Prior to our study, MNPs have been detected in human tissues, including the placenta, but in very low quantities, often fewer than 1 MNP per gram or cm^3^ (Amereh et al., 2022; Ragusa et al., 2021). Current detection methods, such as Raman and FTIR microscopy, have significant challenges, particularly in detecting MNPs in the lower size range, which are considered more hazardous (Dzierżyński et al., 2024). These methods also involve lengthy sample preparation processes, including chemical dissolution and filtration, which inherently lead to the loss of histological information. Additionally, they carry a high risk of external sample contamination, as well as MNP degradation and alteration, compromising the accuracy and reliability of the results (Renner et al., 2018).

*Added value of this study:* This study introduces a universal analytical workflow for MNPs detection in tissues that addresses critical limitations of existing methods. By eliminating external contamination risks and enabling the detection of significantly smaller MNPs, this method revealed orders of magnitude higher MNPs loads in human placenta samples compared to previous reports. The preservation of histological information allows for detailed spatial mapping of MNPs, contributing to a more comprehensive understanding of their distribution and potential biological impacts. A key finding of this study is the positive association between MNP load and placental DNA damage, as quantified by γ-H2AX labeling. This direct correlation between MNP exposure and genotoxic effects strongly suggests that the observed DNA damage is not an artifact of external contamination but rather a consequence of internalized MNPs within the tissue. This first finding of the presence of γ-H2AX foci, a well-established biomarker of DNA double-strand breaks, underscores the potential genotoxic risk posed by MNPs and highlights their ability to induce cellular harm at the molecular level.

*Implications of all the available evidence:* Our research indicates that the true MNPs load in human tissues may be significantly higher than previously recognized. The analytical workflow presented here has the potential to transform the field of MNPs research by enabling more accurate assessments of MNP prevalence in human and other biological tissues. Designed with methods and techniques widely available to researchers across disciplines, this workflow can be readily applied to diverse biological and environmental studies. This, in turn, could inform future studies on their biological interactions, long-term health effects, and dose-response relationships. Given the widespread presence of MNPs in the environment, our findings underscore the urgent need for longitudinal studies to evaluate the chronic effects of exposure, particularly in vulnerable populations such as pregnant individuals and their developing fetuses. Additionally, our findings and methodology are a stepping stone towards true direct toxicological research on MNPs. Addressing MNP contamination at the source and improving public and regulatory awareness are critical steps toward developing effective mitigation strategies.

## Introduction

The pervasive presence of MNPs in the environment and their detection in human tissues have raised significant concerns about potential health impacts (Camerano Spelta Rapini et al., 2024; Donisi et al., 2024; Dzierżyński et al., 2024). Although several studies have reported MNPs in human tissues using workflows typically involving digestion, filtration, and subsequent analysis with FTIR or Raman microspectroscopy (see Table 1), a universally accepted methodology to quantify MNPs in tissues with high sensitivity and spatial precision remains lacking.

**Table 1:**
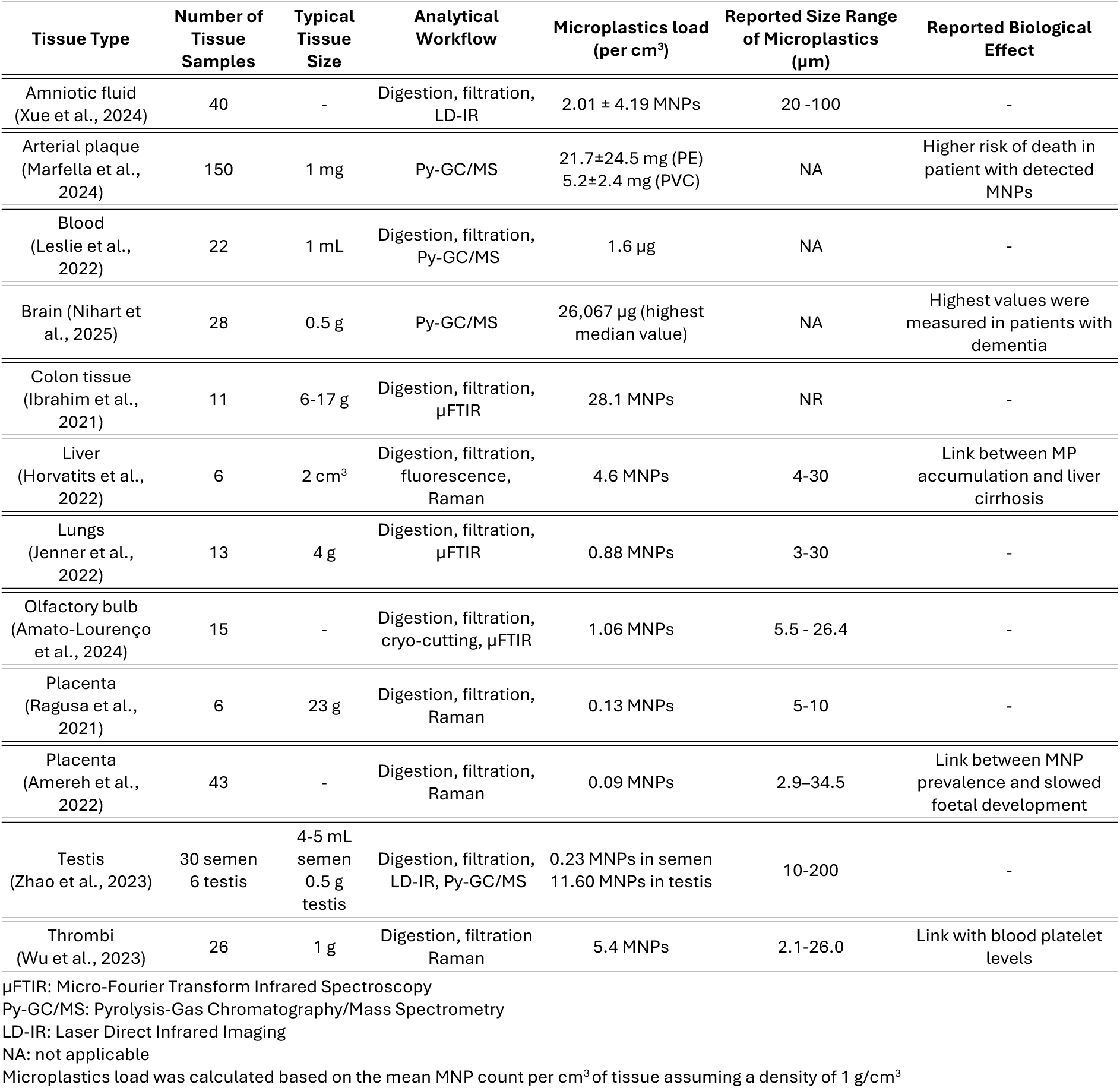
Reports of micro- and nanoplastics (MNPs) detected in human tissues. This table summarizes studies reporting MNPs in various human tissues, detailing sample sizes, analytical workflow used, detected MNP loads, size ranges, and observed biological effects.

Current detection techniques, including Pyrolysis-Gas Chromatography Mass Spectrometry (Py-GCMS) and vibrational microspectroscopy, face considerable challenges. While Py-GCMS quantifies the total plastic load, it does not provide spatial information or retain histological context. In contrast, FTIR and Raman microscopy provide size-related information and have spatial mapping capbilities. However, these methods struggle to detect smaller, more biologically active MNPs, which are often under 5 µm in size, and require lengthy, complex sample preparation, involving chemical dissolution and filtration. Such processes can degrade or alter MNPs, increase contamination risk, and inherently result in the loss of critical histological information (Renner et al., 2018; Van Raamsdonk et al., 2020).

Given these limitations, there is an urgent need for a widely accessible methodology capable of accurately detecting MNPs in tissues from natural exposure. Such a workflow should preserve the histological blueprint and biologically relevant information while being applicable to sufficiently large sample volumes for statistical relevance. It should also enable high throughput analysis, minimize the risk of external contamination from the air, chemicals, solvents, and labware, and possess high sensitivity towards the smallest MNPs, due to their elevated biological activity and diffusivity. Additionally, to support standardization in the field, the methodology should be based on widely accessible, reproducible procedures and analytical methods.

In this study, we develop and validate a universal method for detecting MNPs in tissues, using hydrogel-based tissue transformation and optical clearing combined with MNP staining and readout by fluorescence microscopy. This workflow preserves the histological blueprint of tissues by chemically and enzymatically clearing the sample of interfering materials using mild treatments, while leaving MNPs embedded in their original spatial context within a now background-free hydrogel matrix. External contamination, a frequent issue in MNP research, is effectively prevented as foreign MNPs cannot penetrate the intact or hydrogel-transformed tissue. Fluorescent staining of MNP with Nile Red (NR) is central to this workflow, as it (1) enables detection of MNPs an order of magnitude smaller than current methods, (2) offers significant analytical processing gains needed for future high-throughput analysis, and (3) leverages the widespread availability of fluorescence microscopy.

This validated workflow allowed for a comprehensive analysis of placental tissue, revealing MNPs at previously undetectable sizes and concentrations, with quantities reaching orders of magnitude higher than previously reported (table 1), ranging from thousands to millions per cm³. Furthermore, we found a correlation between MNP load and observed DNA damage in placental tissues, as indicated by labeling of the γ-H2AX histone. These findings underscore the transformative potential of developed approach for future large-scale toxicological research on MNP exposure and associated health impacts, such as immune-inflammatory responses, oxidative stress, accelerated aging, endocrine disruption, and developmental outcomes (Chartres et al., 2024; Donisi et al., 2024; Yong et al., 2020). By establishing a sensitive, reproducible, and contamination-free workflow, we lay the groundwork for advancing our understanding of MNP-related health perturbations in human tissues with unprecedented resolution and reliability.

## Methods

### Sample collection

The ENVIR*ON*AGE (ENVIRmental influence *ON* AGEinG in early life) birth cohort study is an ongoing population-based prospective study recruiting pregnant women giving birth in the East-Limburg Hospital (ZOL; Genk, Belgium) (Janssen et al., 2017a). Mothers were asked to fill out a questionnaire to obtain lifestyle information. Women who were able to complete questionnaires in Dutch, provide written consent, and without a planned caesarean section were considered eligible for participation in the ENVIR*ON*AGE birth cohort study. Because cigarette smoke is a source of DNA damage, we excluded participating mothers who reported that they ever smoked. The study was approved by the Ethics Committee of Hasselt University and East-Limburg Hospital. Under the ENVIRONAGE study, clinical and lifestyle data is collected on the mother-newborn pairs, with additional follow-ups throughout the developmental years of the child. Tissue samples of approximately 100 mm^3^ were derived from the placenta at 4 standard sites near the chorionic plate of the placenta across the middle region, approximately 4 cm away from the umbilical cord and under the chorion-amniotic membrane (Janssen et al., 2017b, Supplementary). Within 10 minutes of delivery, the placentae biopsies were deep-frozen first at −20 °C and after 24 hours at −80 °C until further processing. A randomized and anonymized set of 10 placental samples (delivery dates between 2015 and 2019) was selected for analysis. These samples were used to develop the Standard Operating Procedure (SOP). To ensure objectivity and prevent researcher bias, the MNP measurements were performed blinded to the DNA damage results, were withheld until after the quantification of MNP loads was completed.

### Tissue hydrogel transformation

Tissue coupes with a thickness of approximately 500 µm were cut from frozen tissue cubes. Due to the nature of the gelation protocol, allowing only for molecular transport, and the fact that only the center of the tissue coupes was imaged, complex methodologies for avoiding microplastic contamination could be minimized post-gelation. Pre-gelation, contamination risk is excpected to be minimal, as molecular diffusion (i.e. the gel solution) is very slow, indicating the unlikeliness of any contaminations to penetrate into the core of the tissue coupe.

The samples were incubated overnight with a 0.1 mg/mL Acryloyl-X in 1x PBS, followed by two 15-minute washing steps with 1 mL of 1x PBS. A gelation chamber was constructed out of two cover glasses with a 1 cm gap placed on a Sigmacote-treated glass slide. A total of 200 µL of gel solution was made by adding activation agents 4-hydroxy-TEMPO, TEMED and APS to a gelation stock [0.915 M sodium acrylate; 0.352 M acrylamide; 0.1 M N, N’-methylenebisacrylamide; 2 M NaCl; in 1x PBS] and placed in the chamber.

For optimal hydrogel-monomer diffusion throughout the tissue slice, 100 µL of gel solution was applied to the tissue for 15 minutes at 4 °C. The tissues were transferred to the gelation chamber, and the chamber was sealed with a top coverglass. Incubation proceeded for 2 hours in a pure nitrogen environment at 37 °C. The hydrogel-transformed tissue was then cut to the desired size and region. Over the course of the digestion and labeling steps, the hydrogel swells and expands isotropically. This improves the resolution through physical expansion by a factor of 2 to 4 times the original dimensions. MNPs are non-expandable and remain in the same location respective to the context.

To effectively remove all lipids, which can interfere with the MNP detection, a lipase digestion step was introduced. The hydrogeled-transformed samples were incubated with 1.5 mL of a 10 mg/mL lipase [Sigma Aldrich, 200 U/g] in digestion buffer [0.05 M TRIS; 1 mM EDTA; 2.5% Triton X-100; 0.5 M NaCl] overnight at 37 °C. This was followed by two 5-minute washing steps with 1 mL of 1x PBS to remove remaining enzyme. To further break down remaining proteins, a proteinase K digestion was carried out. The gels were incubated with 1.5 mL of a 10 µL/mL proteinase K [New England Biolabs, 800 U/mL] in the digestion buffer overnight at 50 °C, followed by another two washing steps with 1 mL of 1x PBS.

Biological context labeling was performed using the phalloidin-derived Actin ExM 405 [Chrometra, Belgium] label for actin or the Membrane ExM 405 [Chrometra, Belgium] label for membrane labeling (Wen, 2022). Prior to tissue hydrogelation, the tissue coupes were permeabilized with 1 mL of 0.5% Triton X-100 in 1x PBS for 15 minutes at room temperature to enhance label penetration. For actin labeling, 200 µL of a 0.25 µM Actin ExM 405 solution in 1× PBS was added, while 50 µL of the 0.25 µM Membrane ExM 405 label was used for membrane labeling. For the MNP load quantification experiments, cell nuclei were briefly stained with 1 mL of 1 µg/mL DAPI in 1x PBS.

### Nile Red staining of MNPs

Apolar solvents commonly used for NR-based microplastic staining induce extreme shrinking of the hydrogel matrix and have the potential to swell or dissolve plastics. To resolve this incompatibility issue with our technique, NR was first dissolved in Tween-20 to form a 0.1 mg/mL stock solution was used to form a micellar suspension (Park et al., 2022). The MNPs retained in the hydrogel-transformed tissue were stained with 1mL of a dilute 1 µg/mL NR-Tween-20 in 1x PBS solution at 70 °C for 1 hour. These elevated temperatures are used to loosen the plastic polymer matrix to allow NR to embed itself in the plastic matrix. The gel was washed with 1mL of 1% TWEEN-20 in 1x PBS and again with 1 mL of 1x PBS to remove any residual NR that is not associated with the plastics from the gel before imaging.

### Re-embedding of the hydrogels

After context labeling and MNP staining, the optically cleared hydrogels were re-embedded by cross-linking a secondary, non-expandable gel into the primary hydrogel and onto a microscopy slide. This pocess stops any expansion or contraction of the gels and enables stable imaging This was achieved by treating the slide surface with bind-silane (Cytiva, US) to enable gel fixation. Re-embedding gel solution [0.422 M acrylamide; 0.1 M N, N’-methylenebisacrylamide; 5 mM TRIS] was activated by adding 4-hydroxy-TEMPO, TEMED and APS and used to flush primary cleared gel 2 times for 10 minutes in the fridge, followed by incubation for 1.5 hours in a pure nitrogen environment at 37 °C. Gels were stored in 1x PBS at 4 °C. During hydrogel transformation, the tissue isotropically expanded approximately 3-fold, which remains fixed after re-embedding.

### Fluorescence microimaging and MNP load determination in hydrogel-transformed tissues

A Leica Microsystems TCS SP8X microscope (Leica Microsystems GmbH) with a 25x water immersion objective (NA 0.95) was used to acquire the fluorescence microscopy images across multiple channels and fluorescence lifetime values. Excitation was performed using a supercontinuum White Light Laser at 405 nm for DAPI/Actin ExM 405 and at 488 nm for NR. Emission was captured with hybrid HyD SMD detectors from 415 to 480 nm for DAPI/Actin ExM 405 and from 500 nm to 700 nm for NR in sequential mode. Simultaneously, optical transmission images were recorded using a PMT. Images were acquired with 1024 x 1024 pixels at 400 Hz scan speed and 4-line accumulations. For particle counting, re-embedded hydrogel-transformed tissues were subjected to a large volume fluorescence imaging approach comprising of a 5×5 tile scan covering a total area of 1 mm by 1 mm with a thickness of 100 µm, for a total imaged volume of 0.1 mm^3^ with a pixel size of 200 nm. Each stack consisted of 100 z-slices, which were stitched together in the LAS X software to reconstruct the full 3D volume. To mitigate the risk of recording external MNP contamination, the imaged volumes were selected at least 50 µm below the surface of the hydrogel-transformed tissues.

MNPs were counted automatically using a custom software tailored to the large size of the imaged volumes. The particle counter employs a threefold identification strategy for extracting particle counts from the stitched images. Firstly, the intensity values in the NR channel were thresholded by extracting only the 99.97’th percentile of the highest intensity voxels. This step effectively removes low-intensity background fluorescence and residual artefacts from the data, ensuring that only the brightest regions, most likely corresponding to MNPs, are retained for analysis. Next, the resulting thresholded images were further refined by selecting only voxels with a prominence of at least five intensity units above the surrounding voxels. This step filters out faint peaks that may result from random noise or diffuse background fluorescence, ensuring that only distinct, well-defined intensity peaks, characteristic for NR-stained MNPs, are counted. Finally, size thresholding was applied to include only voxel clusters comprising at least 12 connected voxels in three dimensions. This size-based filtering excludes small, random intensity specks that could result from noise, and was empirically determined by calibrating the detection system with 400 nm polystyrene particles and measuring the corresponding signal and volume. This approach provided an optimal balance between avoiding MNP overestimation through false positives from artefacts and accurately assessing the real number of particles visualized in the NR images.

**Figure 1.**
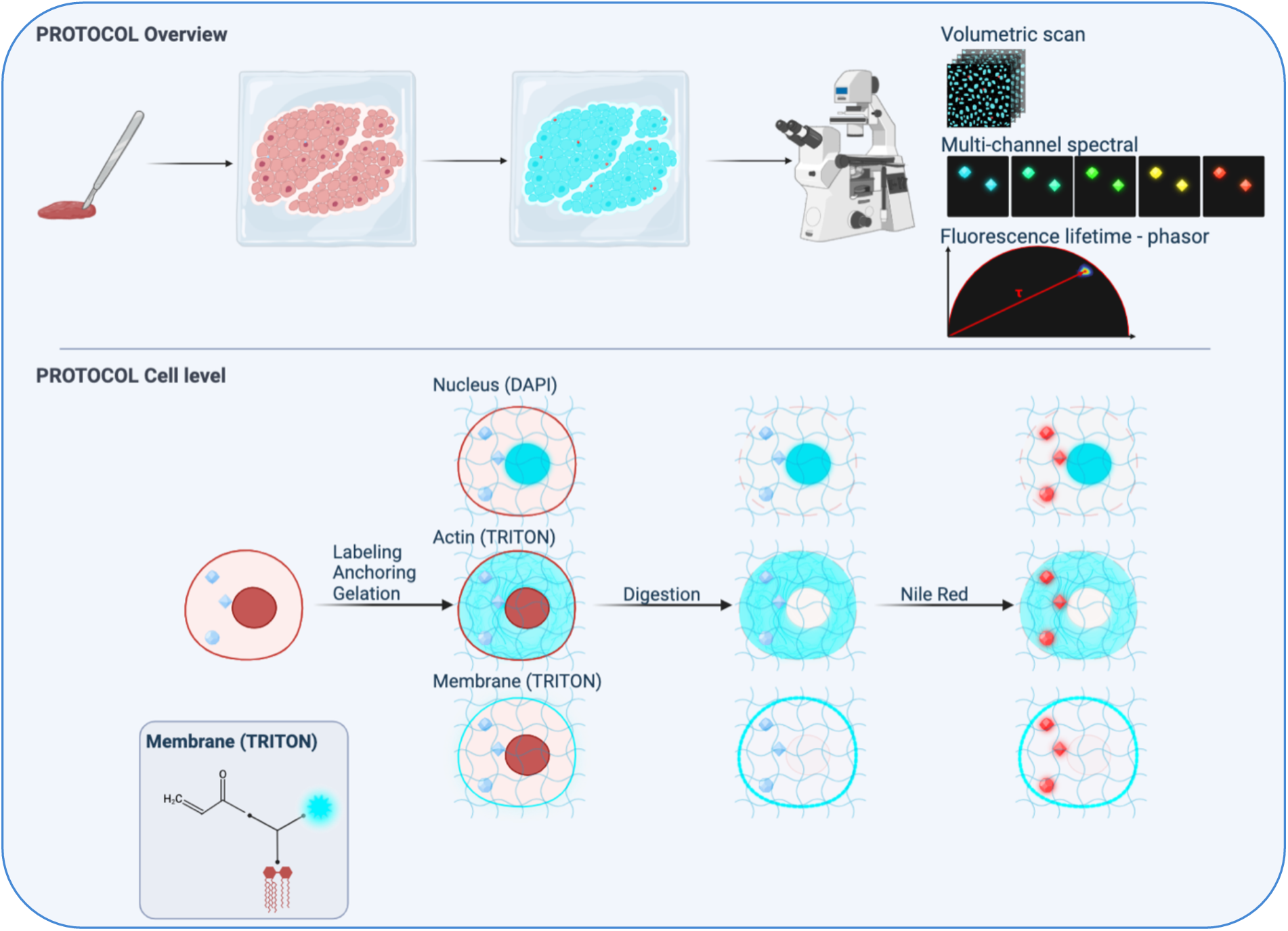
Protocol for microplastic detection in tissues through optical clearing. Top: overview of the hydrogel-transformation workflow and multimodal MNP identification. Bottom: relevant biological context is labeled with TRITON labels which are cross-linked to the gel. Proteins are also chemically linked to the gel, fixing the bio-structure in place. A two-fold digestion with lipase and proteinase removes all biological structures and potential artefacts. Labels from the relevant context and MNPs remain fixed in the gel. Finally, optimized NR staining enables localization and identification of the MNPs in the sample.

### Spectral and Fluorescence Lifetime based MNP identification

Identification of the chemical nature of MNPs was performed by assessing both the fluorescence spectral and temporal responses of NR-stained structures. The fluorescence lifetime of NR serves as a reporter for the surrounding environment, with distinct lifetimes when embedded in lipids compared to the most common plastics (S4) (Sancataldo et al., 2020). Lifetime measurements were acquired using fluorescence lifetime imaging (FLIM). NR lifetime estimations were obtained by taking 50 accumulations of time-correlated single photon counting (TCSPC) of each image area using a femtosecond pulsed laser with a repetition rate of 80 MHz. The laser intensity was adjusted to achieve a total of at least one million photon counts per acquisition to consolidate the lifetime estimation. Rather than using model-based tail fitting, the phasor approach was used to evaluate lifetimes. A phasor plot grouping the fluorescence decay data for each pixel enabled rapid lifetime estimation and visual identification of polymer types (Zhou et al., 2022). FLIM measurements served two purposes: (1) to validate the clearing efficiency by ensuring that lipid signatures were no longer detectable post-digestion, thus preventing false positives, and (2) to classify the plastic type on the phasor cloud distributions (Figure S3).

For the spectral response, a solvatochromatic shift in the emission spectrum of NR is expected, depending on the chemical nature of the environment (Meyers et al., 2022). This shift was detected using a multi-channel approach, with 5 detectors for simultaneous capturing of the fluorescence signal in 40 nm intervals starting at 500 nm up to 700 nm, labeled Ch1 to Ch5 (Leica TCS SP8X with 3 PMTs and 2 HyD detectors; Figure S2). These channels were used to generate the ratiometric intensity parameters (i.e. Ch1/Ch2, Ch/Ch3, Ch3/Ch4, Ch4/Ch5 and Ch5/Ch1) used to classify the plastic types. Classification was performed using an decision tree model that was optimized in Classification Learner in MATLAB. The model was trained using the ratiometric values for 50 particles originating from cryomilled plastic pellets (Retsch Cryomill; PP, PE, PVC, PET, PS) to define the decision tree. Class separation at each decision node was optimized using the Gini diversity index and the parameters were optimized using 30 Bayesian iterations. Model performance was evaluated using the minimum classification error (MCE), accuracy, sensitivity (TPR) and specificity (FNR). The trained model used for identification achieved 91% accuracy. A confusion matrix (Figure S4) was constructed to reflect the model’s performance in categorizing particles.

### DNA damage quantification

The unprecedented levels of MNPs detected in these placentas raise critical concerns about their biological impact, particularly their ability to induce DNA damage. Using fluorescent immunolabeling of the γH2AX marker for DNA double strand breaks (Mah et al., 2010), we quantified DNA damage as a sensitive parameter of genotoxic stress. The γH2AX parameter, calculated as the ratio between the γH2AX-foci count and the nuclei count, provided a robust measure of DNA damage. Counts were averaged over ten sections per sample, with imaging performed and analyzed in ImageJ. The MNP load was logarithmically transformed (log10) to meet the linearity assumptions. We computed correlation coefficients between the DNA damage and the MNP load by applying both parametric (Pearson) and nonparametric (Spearman rank) correlation coefficients. In the partial correlation models (both parametric and nonparametric), we adjusted for the following covariates and confounders: newborn sex, gestational age, birth parameters (length and weight) and maternal age.

## Results

### Study population characteristics

For this study, randomly selected placental biopsies of 10 mother-newborn pairs including six girls and four boys were used. Median birth weight was 3.405 kg (range: 3.210-3.630), median birth length was 49.75 cm (range: 48-52), and the median gestational age was 281.5 days (range: 274-284). The median maternal age was 29.5 years (range: 26-37), and the average pre-pregnancy BMI of the mothers was 23.5 (range 18.4-28.7). All mothers were non-smoking, and six out of ten mothers were ethnic Europeans, while four had non-European origins (defined as having at least three grandparents of non-European descent).

### Development of tissue hydrogel transformation for reliable MNP detection

Vibrational microspectroscopy techniques, including IR and Raman spectroscopy, have shown limitations in the detection of the most biologically relevant MNPs. These methods generally lack the sensitivity to reliably detect particles smaller than 5 µm and are time-intensive, making them unsuitable for high-throughput analysis. In contrast, fluorescence microscopy, particularly when combined with selective MNP staining, presents a highly sensitive alternative, albeit currently primarily used to rapidly screen larger microplastics. We demonstrate that NR staining enables routine detection of MNPs smaller than 1 µm and allows differentiation based on distinct fluorescence properties (Figure S5).

A critical challenge in MNP detection with NR staining is the risk of false positives due to co-staining of apolar structures, such as lipids, within tissues (Meyers et al., 2022). To address this, we implemented hydrogel-based tissue transformation, which removes interfering biological components that would otherwise be NR-stained and misinterpreted as MNPs, while embedding the tissue’s structural blueprint into the hydrogel matrix. This transformation still allows complementary selective fluorescence labeling of biological structures of interest, providing relevant histological context within the hydrogel-transformed tissue, while preserving MNPs in their original positions and enabling fluorescence-based identification.

To ensure that residual traces of lipids and other biomolecules, which would lead to false positives, are completely removed, we developed a two-step enzymatic digestion protocol at elevated temperatures, combined with extensive gel washing steps using concentrated surfactants. The optimized protocol was validated using cell spheroids grown with 3 µm and 400 nm polystyrene (PS) MNPs intentionally added. These cell spheroids serve as a reproducible 3D biological model due to their uniform size and structure. The experiments demonstrated that the MNPs remained embedded throughout the hydrogel matrix and were effectively stained and detected within the entire 3D volume (Figure S1). The hydrogel transformation also provides optical clearing, aiding signal detection across the full depth of the sample, and confirming that MNPs below 1 µm can be easily detected througout (Figure S1). Lipid structures were fully removed, as no residual NR staining was observed post-clearing (Figure 2, Figure S1), preventing false positives from co-stained apolar components. Importantly, the validation demonstrated that MNP retention within the hydrogel matrix remained consistent, with no changes in MNP count, fluorescence intensity, or localization within the 3D volume over time.

Furthermore, the optical homogenization of the hydrogel matrix also enables MNP visualization through regular optical transmission imaging (Figure 2B, Figure S1B,D), providing an independent validator of NR fluorescence signals. Given that the sizes of the suspected MNPs did not allow for identification using Raman spectroscopy, machine-learning mediated spectral analysis (Figure S6-8) and lifetime imaging were used to enable characterisation of the particles.

### Validation in placental tissue

Further validation on placental tissue slices (500 µm thick) demonstrated that the hydrogelation procedure was effective in more complex tissue samples, despite their increased biological heterogeneity. Placental tissue contains heterogeneous structures, such as varying cell types, extracellular matrix, and blood vessels, which can introduce diffusion barriers during enzymatic digestion and clearing. Our optimized protocol effectively removed interfering biomolecules, such as lipids and other apolar components that could otherwise be misstained with NR, while retaining MNPs throughout the entire tissue volume (Figure 2A). While NR co-staining of residual tissue material is typically observed with standard hydrogel transformation procedures, no such artifacts were observed following our optimized digestion steps (Figure S1-2). Image analysis reveals a reduction of NR-stained biological structures to below the detection limit under the standardized imaging circumstances in which the MNPs are detected. Finally, even if minimal traces of lipids remain, they can be distinguished from NR-stained MNPs based on their distinct fluorescence spectra and shorter fluorescence lifetimes (NR-stained biological structures: 2,039–2,629 ns vs. > 3 ns for all tested plastics, Figure S4). NR fluorescence spectra and lifetime measurements of relevant plastics, both with and without undergoing the full hydrogel transformation process, confirm that the procedure does not interfere with their fluorescence based detection and identification. The detected particle in Figure 2 exhibits an estimated fluorescence lifetime of 3,668 ns, consistent with our library of NR stained reference plastics (Figure S3, S4), by which it was classified as PVC.

### MNP load determination in placental samples

To assess the prevalence of MNPs in placental tissue, we applied the developed workflow to biopsy samples from 10 different human placentas. From each placenta, a 500 µm thick section was carefully taken from the central region near the chorionic plate. Figure 2 shows two examples where the tissue blueprint was generated by labeling actin with a Fluor-405 fluorescent marker, providing histological context by delineating cell volumes through cytoskeletal labelling. This fluorescence signal is spectrally complementary to the NR emission, enabling precise localization of NR-stained MNPs within the structural features of the tissue. This demonstrates that MNPs are not only present in the interstitial spaces and blood, but that they are taken up inside the placental cell volume. The optical transmission image of this thick section confirms effective tissue clearing, with a distinct particle visible in the transmission image that coincides precisely with the fluorescence particle detected in the NR channel.

MNPs were detected across all 10 hydrogel-transformed placental tissues, each with an imaged volume of 0,1 mm³. Accounting for the volumetric expansion factor of 3^3^ during the hydrogellation process, this corresponds to an original placental tissue volume of approximately 3,7 x 10⁻³ mm³. In these 10 placental samples, a median of 25 MNPs were found (range 2 - 1056), amounting to a median concentration of 6,75 × 10^6^ MNPs per cm^3^ of tissue (Table 1; Figure S5, S9). The median size of the detected MNPs was found to be 0,76 µm (range 0,44 - 3,7 µm), calculated based on the largest cross-section of the detected voxel cluster in the xy plane.

Notably, 73% of the detected MNPs are plastics smaller than 1 µm in diameter, indicating a size distribution that is skewed towards smaller sized particles (Figure S5). Particles below 400 nm in diameter were excluded, as method validation was performed with 400 nm diameter polystyrene particles, and the particle analysis software excludes signals below this size treshold. No significant difference in size was found between different placental samples (Figure S5).

**Figure 2.**
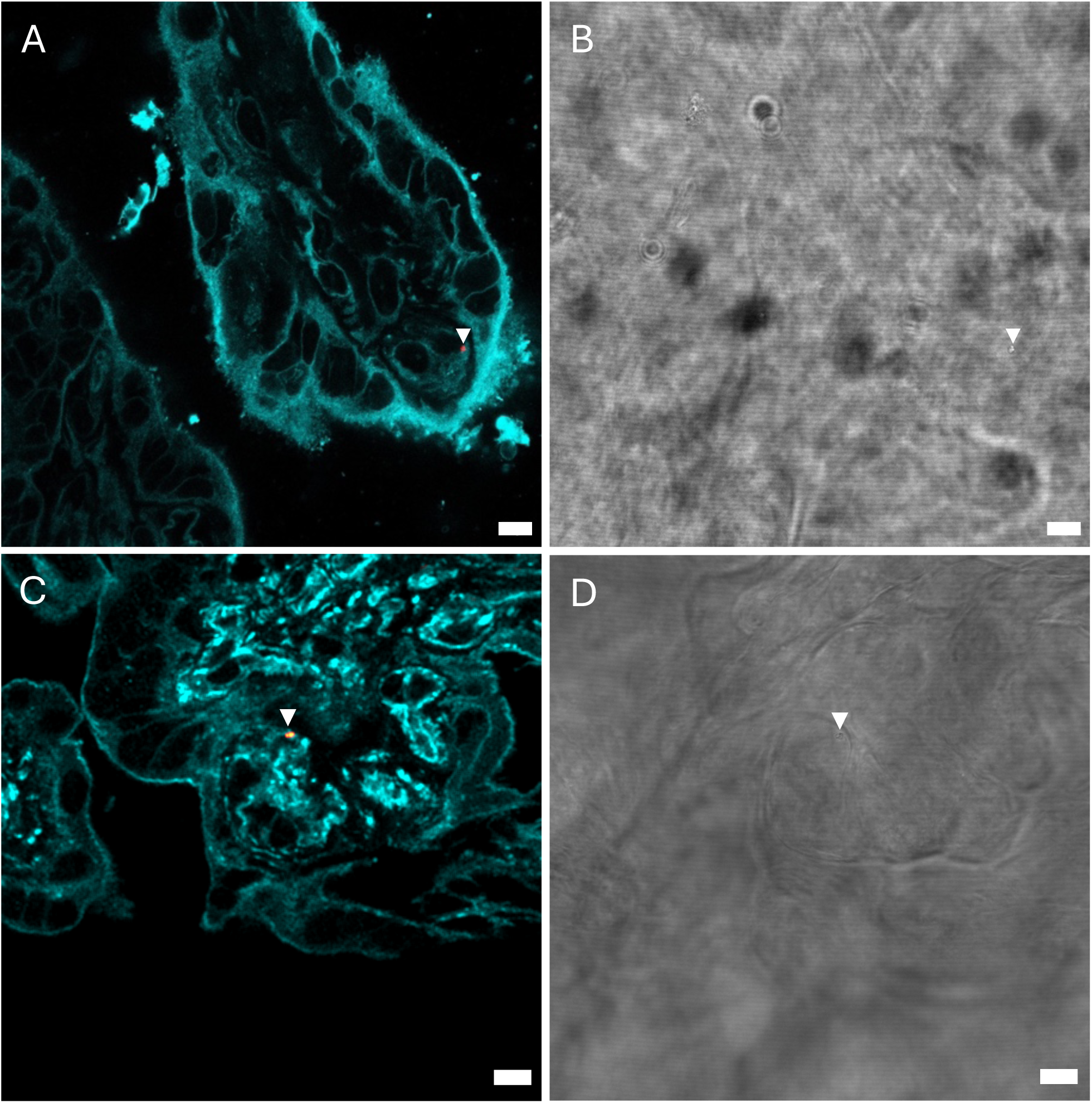
Confocal fluorescence imaging of an optically cleared placental tissue sample with TRITON label for cytoskeletal actin (cyan) and NR stain for MNPs. MNPs (indicated by white triangle) was identified in the NR channel (panel A) and validated via transmission imaging (panel B). Scalebars are 10 µm.

**Table 2.** Detected MNP load across placental tissue samples and corresponding DNA-damage.

| Sample | Absolute particle count | Concentration [MNPs/cm<sup>3</sup>] | Mean diameter [$\mu\text{m}$] | Max diameter [$\mu\text{m}$] | Min diameter [$\mu\text{m}$] | y-H2AX | Standard deviation yH2AX |
| --- | --- | --- | --- | --- | --- | --- | --- |
| 1 | 45 | 12.150.000 | 0,90 | 2,33 | 0,49 | 1,133 | 0,182 |
| 2 | 14 | 3.780.000 | 0,91 | 2,62 | 0,49 | 0,284 | 0,082 |
| 3 | 2 | 540.000 | 0,66 | 0,66 | 0,66 | 0,305 | 0,114 |
| 4 | 27 | 7.290.000 | 0,85 | 3,70 | 0,44 | 0,789 | 0,362 |
| 5 | 24 | 6.480.000 | 0,82 | 1,87 | 0,54 | 0,073 | 0,051 |
| 6 | 1056 | 285.120.000 | 0,87 | 3,20 | 0,38 | 1,169 | 0,155 |
| 7 | 42 | 11.340.000 | 0,93 | 1,66 | 0,44 | 1,059 | 0,498 |
| 8 | 22 | 5.940.000 | 0,97 | 2,77 | 0,49 | 1,000 | 0,264 |
| 9 | 8 | 2.160.000 | 0,77 | 1,46 | 0,49 | 0,697 | 0,380 |
| 10 | 73 | 19.710.000 | 0,85 | 2,10 | 0,49 | 0,162 | 0,081 |

### Correlating MNP load with DNA damage

Placental γH2AX DNA damage averaged 0.67 foci/cell. Before adjustments, as illustrated in Fig. 3, a trend toward a positive association between placental γH2AX DNA damage and the placental MNP concentration was observed (Pearson r=0.47; p =0.17; Spearman rank correlation r=0.43; p=0.20), though this correlation was not statistically significant. The data was then adjusted for key covariates and confounding variables including newborn sex, gestational age, birth length and weight and the age of the mother, leading to a Pearson correlation of r=0.93; p=0.005, which was further supported by the non-parametric Spearman rank partial correlation (r=0.80; p=0.055).

**Figure 3.**
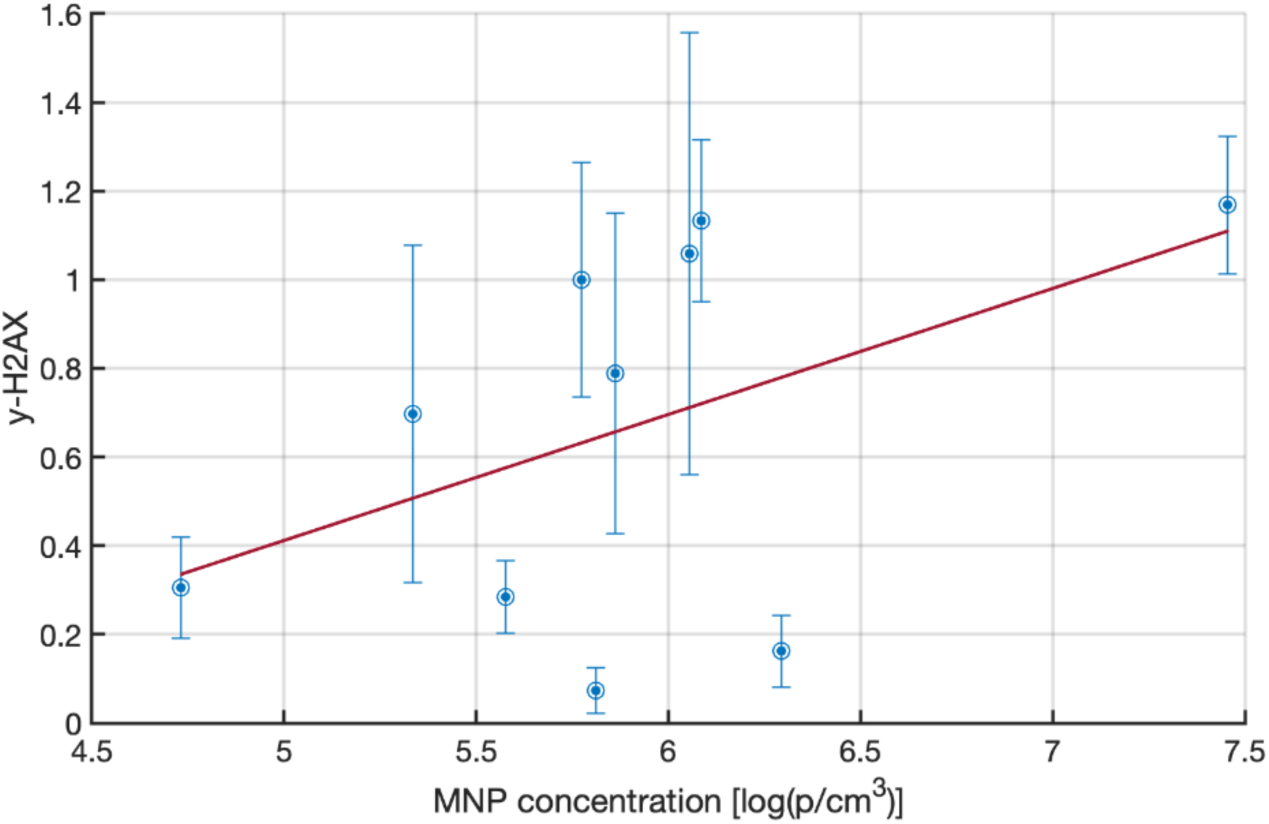
Correlation between measured MNP prevalence in placental tissues and the corresponding DNA damage in the same sample. Linear regression (red line) has a slope of 0.2842 and an intercept of −1.0093. It’s goodness of fit (R^2^) is equal to 0.2196. Whiskers indicate the standard deviation for the γH2AX measurements.

## Discussion

MNPs are pervasive and persistent environmental pollutants that have infiltrated virtually every corner of the globe, including human tissues. Despite their ubiquity, the fate and potential toxicity of MNPs in the human body remain largely underexplored, posing a significant gap in our understanding of their health implications. The placenta, serving as a critical interface between mother and fetus, is particularly vulnerable to such contaminants. According to the Developmental Origins of Health and Disease (DOHaD) theory, prenatal exposures can have profound and lasting effects on health across the lifespan (Lacagnina, 2019). Indeed, prior evidence indicates that environmental particles like black carbon can cross the placental barrier and disrupt fetal development (Bai et al., 2024; Bové et al., 2019). However, accurate assessment of biologically relevant MNPs, especially in the sub-micron range, has been severely hampered by inadequate analytical techniques that fail to preserve tissue architecture and are prone to contamination, limiting our ability to fully comprehend the risks posed by these pollutants.

In this study, we have overcome these significant methodological barriers by developing a groundbreaking hydrogelation-based workflow for the in-situ detection of MNPs. This innovative method preserves histological integrity, thereby maintaining the biological context, while simultaneously minimizing the risk of external contamination—a critical issue in previous studies. Our optimized two-step enzymatic digestion effectively eliminates potential artefacts, facilitating reliable MNP staining with NR and then subsequent precise localization. By integrating this approach with advanced fluorescence microscopy, we have achieved an unprecedented detection resolution, identifying nanoplastics as small as 400 nm. Detection at this scale is particularly crucial given the enhanced biological activity and translocation potential of smaller particles. Moreover, the high-throughput potential of this workflow enables the examination of large tissue volumes, significantly advancing the feasibility of comprehensive MNP analysis in human tissues.

Applying this novel method to human placental samples, we uncovered MNP concentrations that are several orders of magnitude higher than those previously reported, with a median concentration of 6.75 × 10⁶ MNPs per cm³ of tissue. This stark discrepancy indicates that the true MNPs load in human tissues has been drastically underestimated by earlier studies, likely due to methodological limitations. The predominance of MNPs smaller than 1 µm (comprising 73% of detected particles) is particularly alarming, given their capacity to penetrate cellular membranes, traverse biological barriers, and interact intimately with subcellular structures. These findings necessitate an urgent reassessment of environmental exposure levels and raise critical questions about the potential health risks posed by widespread MNP contamination.

Our study also revealed a positive association between MNP concentration and γH2AX immunolabeling, a well-established marker for DNA double-strand breaks, suggesting a potential link between MNP exposure and genotoxic stress within placental tissue. Although the correlation did not reach conventional levels of statistical significance— likely due to our limited sample size—the observed trend is biologically plausible and consistent with the documented genotoxic effects of other environmental pollutants. The amount of MNPs and their presence even within the cell volume, coupled with indicators of DNA damage, raises significant concerns regarding the integrity of placental function. Disruption of placental processes can have profound consequences, as genotoxic stress during pregnancy has been associated with adverse outcomes, causing effects that can persist throughout the child’s life.

By bridging the methodological gap in MNP detection, our study demonstrates the transformative potential of this novel approach for advancing toxicological research. The capacity to detect and quantify MNPs in situ, while retaining tissue integrity, enables a more accurate assessment of their distribution, cellular interactions, and potential biological effects. By correlating MNP load with functional biological markers such as DNA damage, we establish a critical foundation for elucidating the health impacts of these pervasive pollutants. Moreover, this method is broadly applicable and can be adapted to investigate MNP accumulation in other tissues and organ systems, significantly enhancing our ability to map and understand the full scope of human exposure.

Notwithstanding these compelling findings, the current study was conducted on a relatively small sample size, which may have limited the statistical power of our analyses and the ability to detect significant associations. This limitation underscores the necessity for future larger-scale studies to validate our finding. Additionally, while γH2AX is a sensitive and specific marker for DNA double-strand breaks, further research is needed to unravel the mechanistic pathways by which MNP exposure may induce genotoxicity. Investigations into oxidative stress, inflammatory responses, and endocrine disruption are particularly pertinent and could elucidate the multifaceted ways in which MNPs impact cellular function.

In conclusion, our research marks a pivotal advancement in MNP toxicology, shedding light on the previously underestimated prevalence of MNPs in human placental tissue and their potential health implications. By establishing a sensitive, reproducible, and high-throughput detection method, we provide a critical tool that can inform future research, public health initiatives, and regulatory policies. Given the escalating ubiquity of MNPs in the environment and their infiltration into human tissues, there is an urgent imperative to intensify research efforts aimed at understanding and mitigating their health risks. Protecting vulnerable populations, particularly pregnant women and developing fetuses, should be a priority in public health strategies and environmental regulations.

## Data Availability

All data produced in the present study are available upon reasonable request to the authors

## Appendix

**Figure S1.**
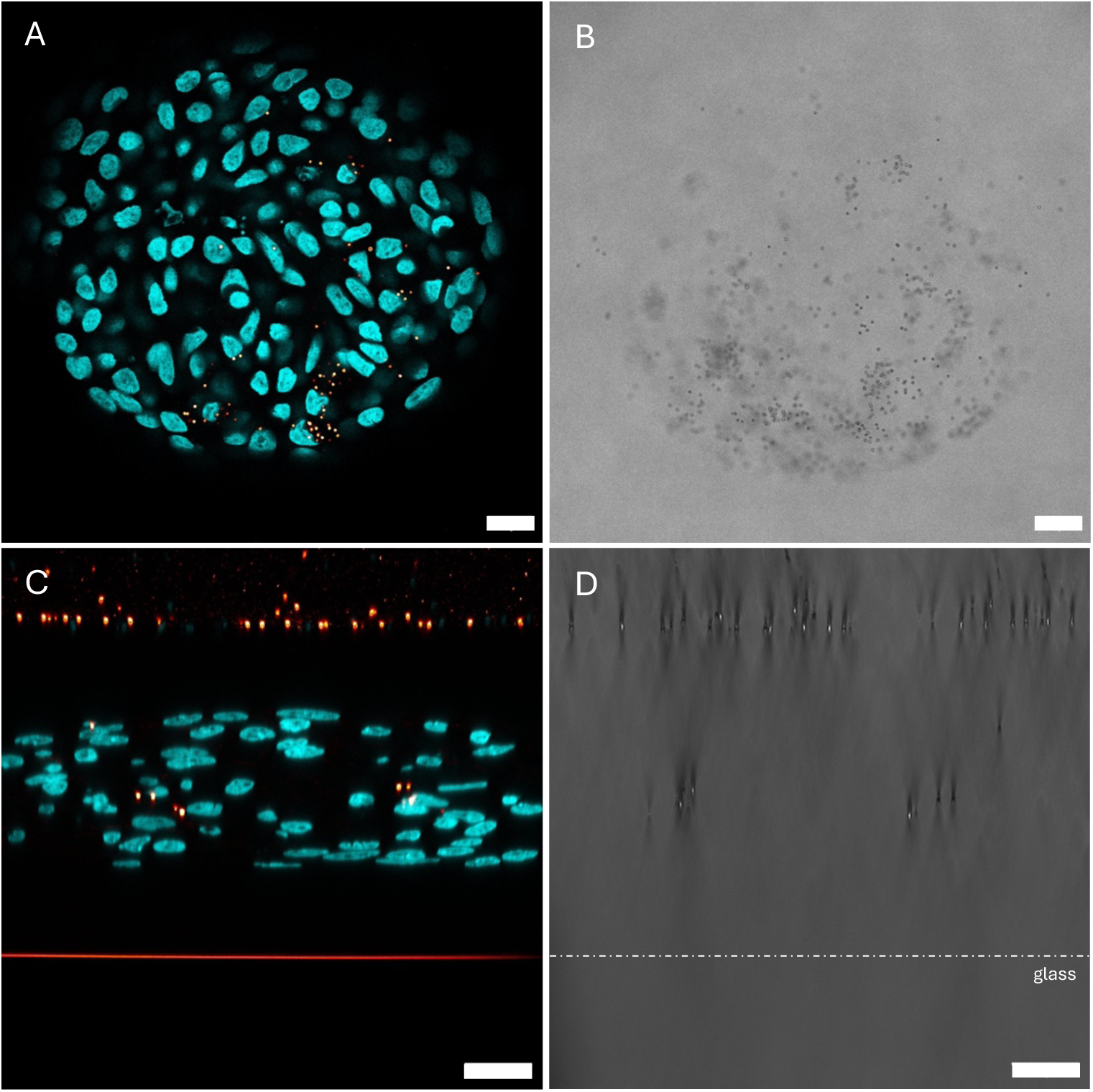
An A549 cell spheroid was cultured in medium contaminated with 3 µm polystyrene particles. The nuclei were labeled with DAPI (cyan) and MNPs with NR (red). In pane A, 3 µm PS is clearly embedded in the spheroid cells, which is confirmed through the transmission image in pane B. No remaining lipid structures can be detected. In panes C and D, an xz cross-section of the gel was taken. A suspension of NR stained 3 µm and 400 nm PS particles was added on top of the gels to simulate contamination events. No particles were detected in the gel between the spheroid and the outside contamination, demonstrating the robustness of the protocol against contamination. Scalebars are 50 µm.

**Figure S2.**
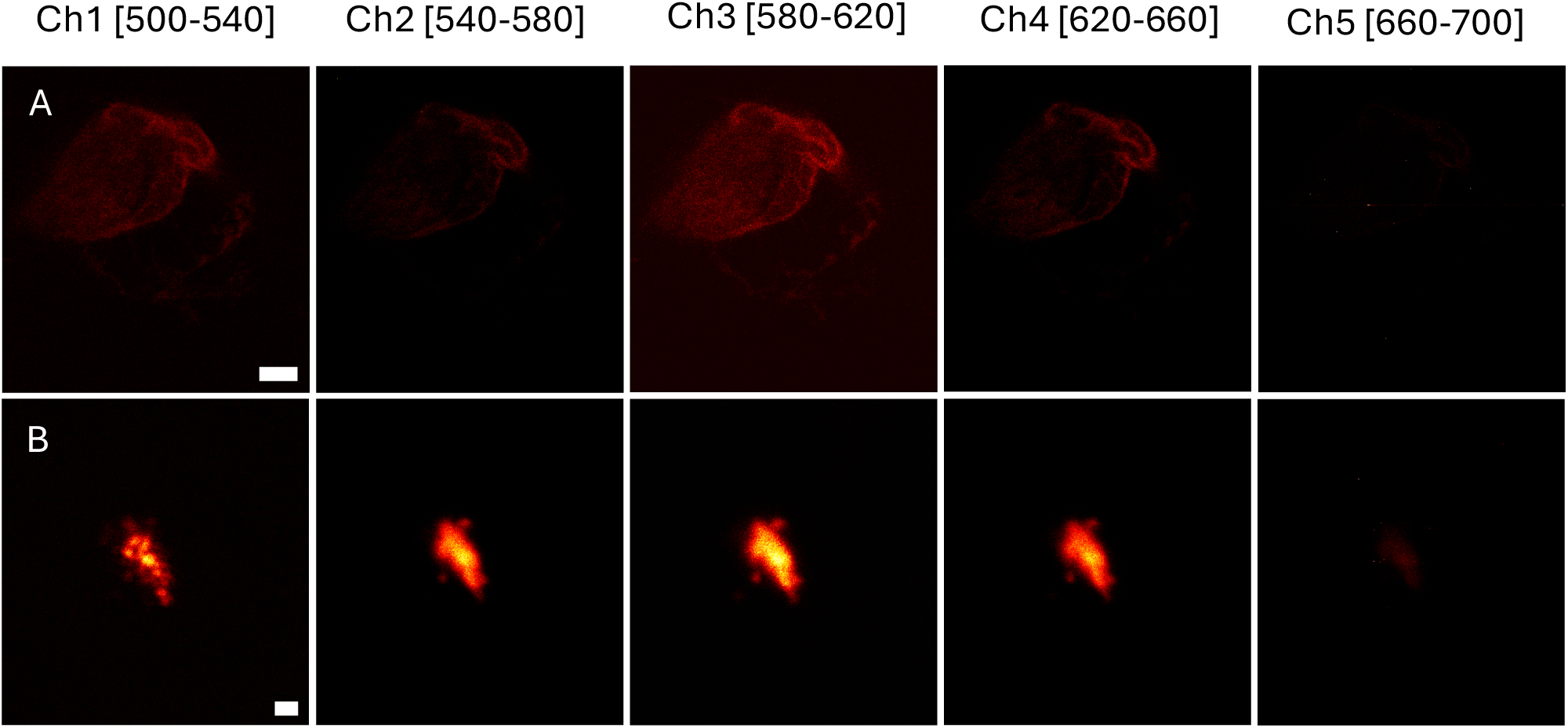
Spectral response of a NR stained lipid artefact (no enzymatic digestion) in row A and of a plastic particle in row B as input for the ratio-metric machine-learning mediated classification. Scalebars are 10 µm in row A and 1 µm in row B.

**Figure S3.**
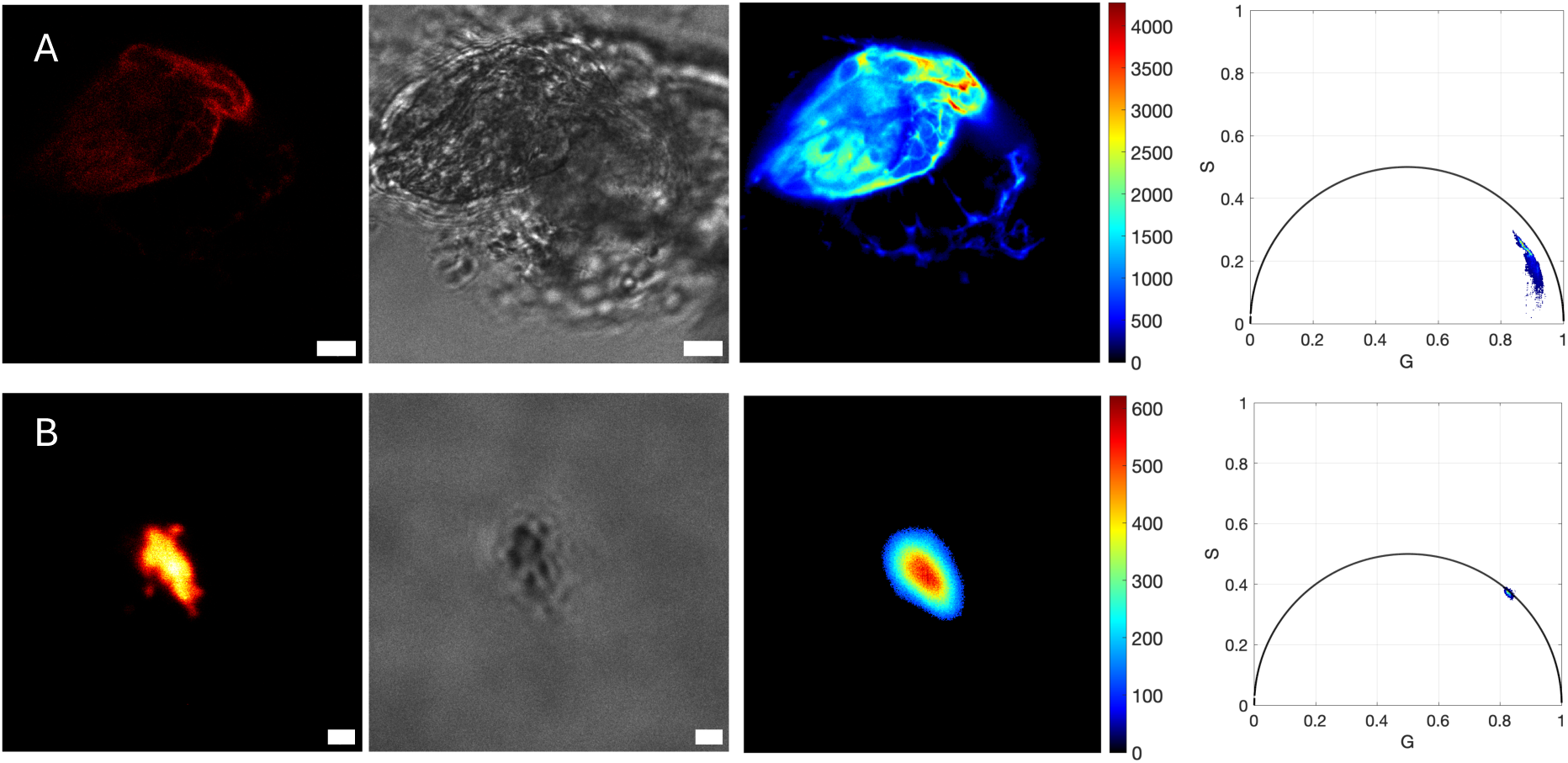
Fluorescence lifetime response of a NR stained lipid artefact (no enzymatic digestion) in row A and of a plastic particle in row B both in a hydrogel. Scalebars are 10 µm in row A and 1 µm in row B.

**Figure S4.**
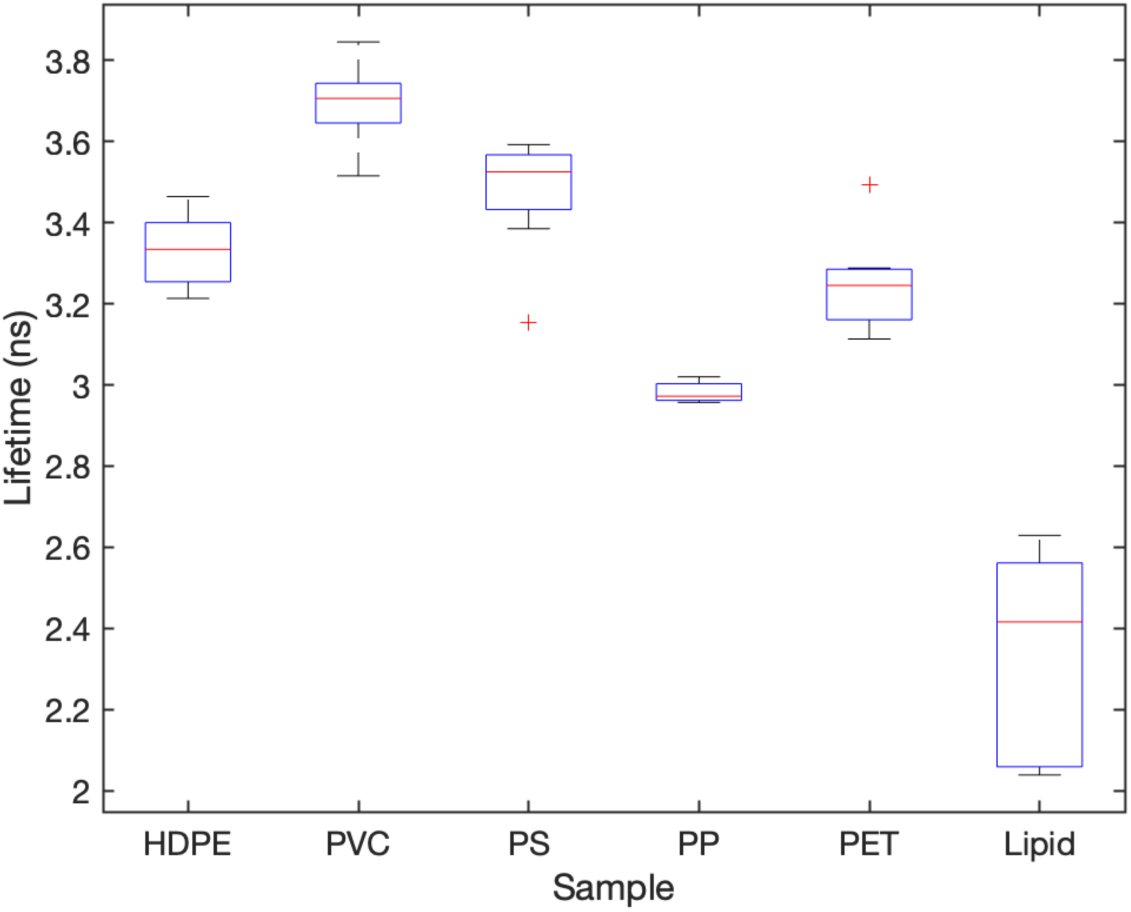
Estimated fluorescence lifetimes for (HD)PE, PVC, PS, PET and PP based on lifetime evaluations of cryomilled reference plastic particles and of lipid structures in hydrogels (n=9).

**Figure S5.**
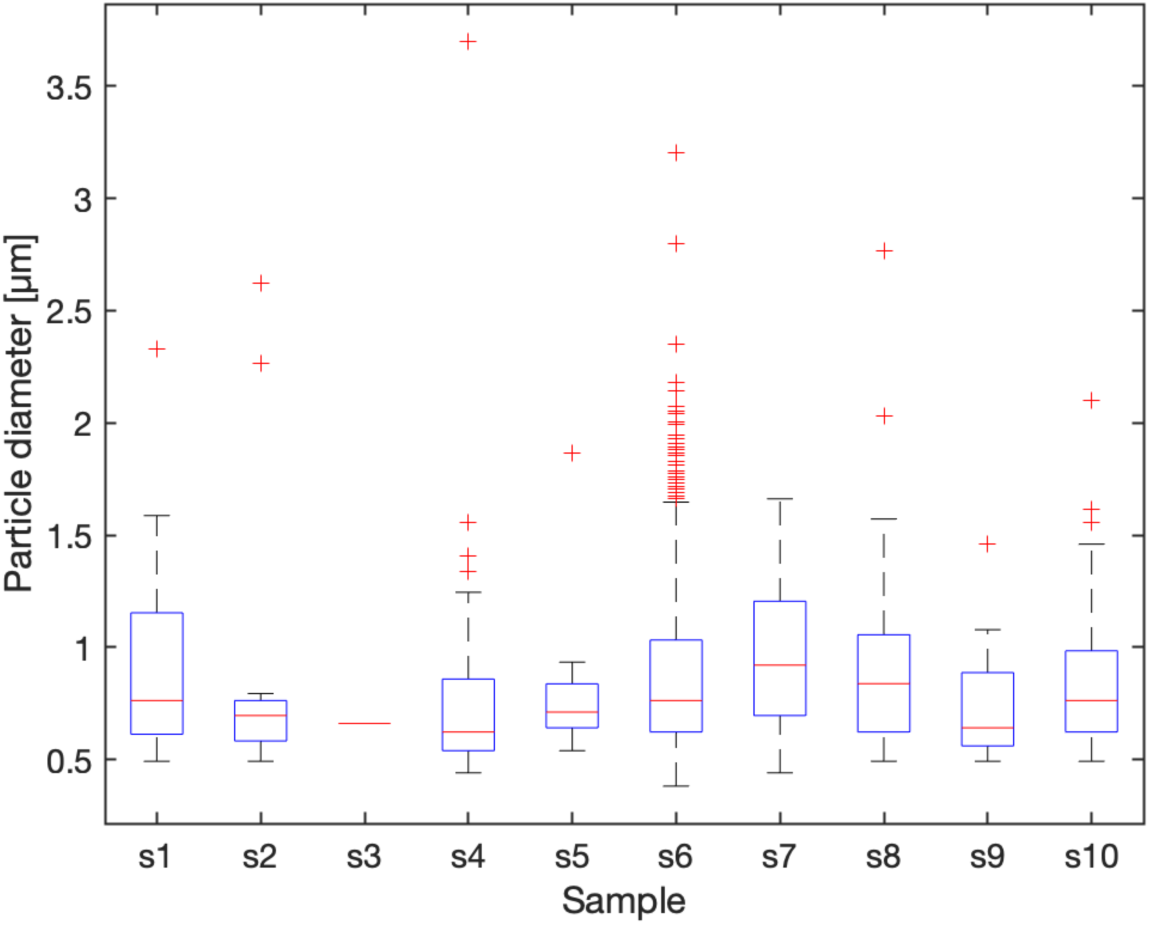
Size distribution of detected MNPs in the placental tissue samples.

**Figure S6.**
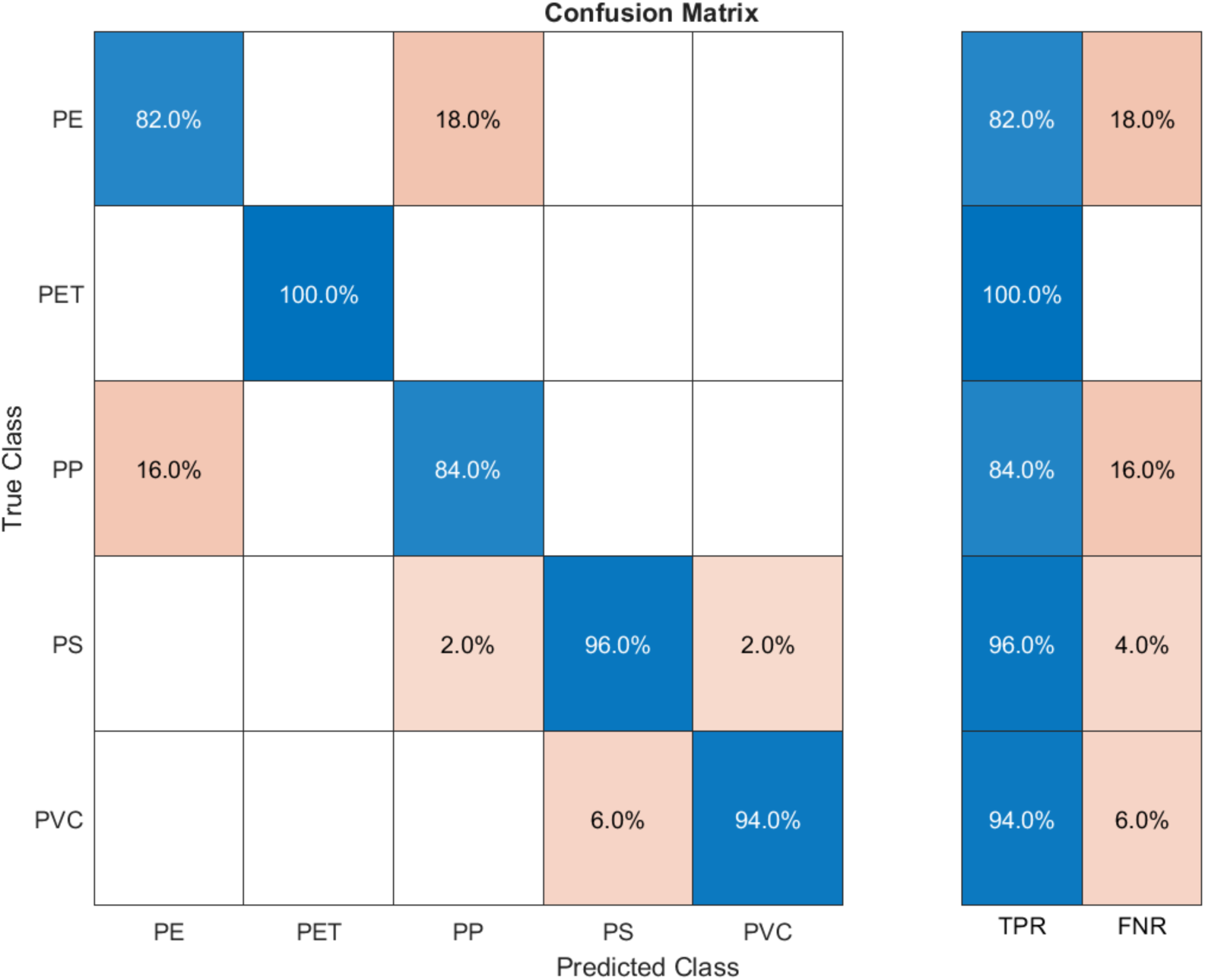
Validation confusion matrix representing the model’s performance, evaluated using 10-fold cross-validation. Only between PP and PE, a slight chance for mismatch was observed. Overall, a true positive rate (TPR) of 91,2% was observed, with a false negative rate of 8,8%. No separate test dataset was used.

**Figure S7.**
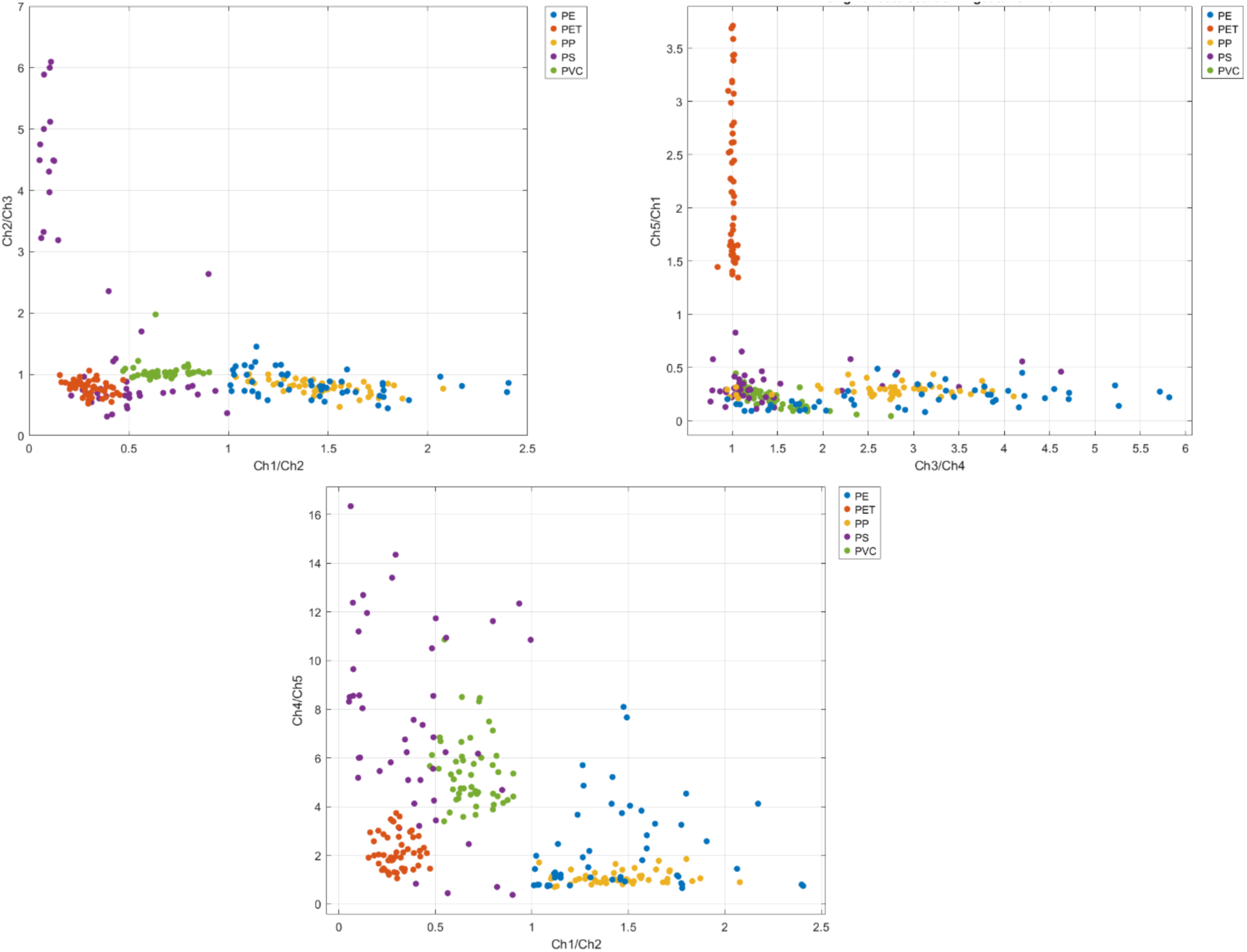
Scatter plots of ratiometric fluorescence intensities for Ch1/Ch2, Ch2/Ch3, Ch3/Ch4, Ch4/Ch5 and Ch5/Ch1 showing clear clustering of plastics, allowing for construction of a decision tree.

**Figure S8.**
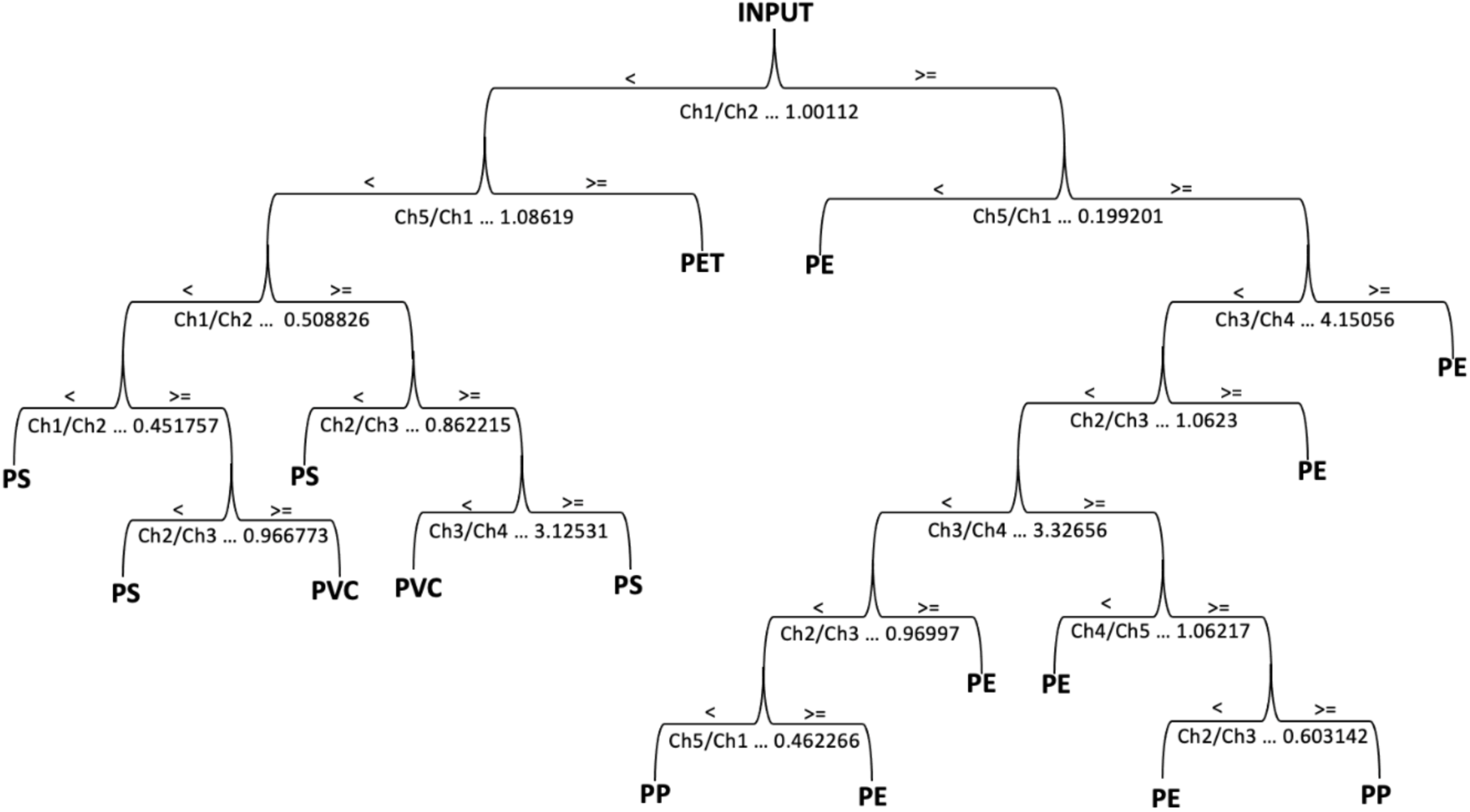
Decision tree indicating optimized decision nodes and ratiometric thresholds for classification of the MNPs.

**Figure S9.**
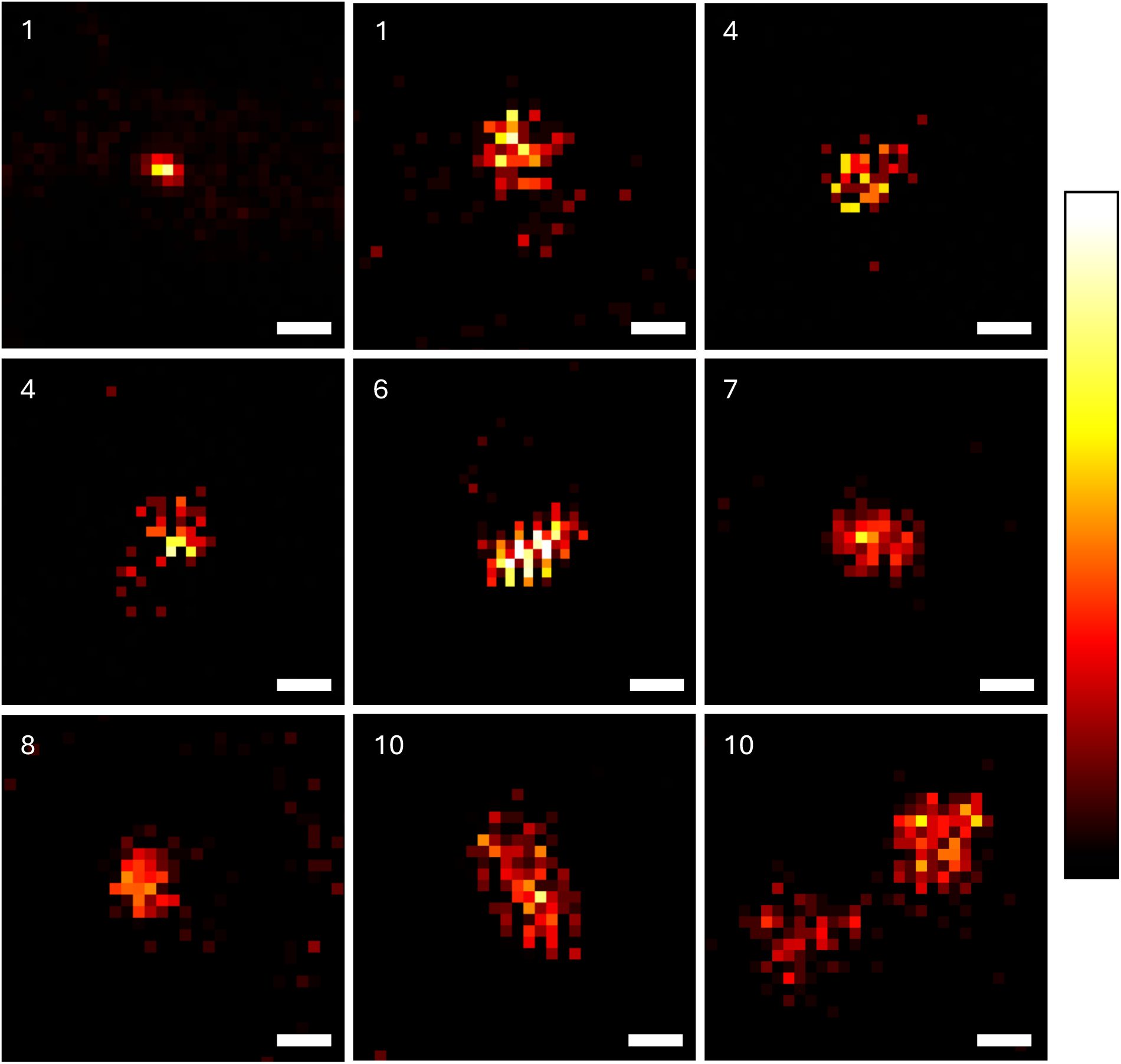
Particles manually identified in various placental samples. The corresponding sample number is indicated in upper left corner. Notably, the signature from these particles only cover a few pixels, even before thresholding and Scalebars are 1 µm.

## Notes

### Competing Interest Statement

The authors have declared no competing interest.

### Funding Statement

This study was funded by research Foundation Flanders - FWO

### Author Declarations

Ethics committee of University Hospitals Leuven gave ethical approval for this work.

